# Incidence and predictors of pulmonary exacerbations in primary ciliary dyskinesia: an international prospective cohort study

**DOI:** 10.1101/2025.01.13.25320478

**Authors:** Leonie D Schreck, Myrofora Goutaki, Eva SL Pedersen, Fiona Copeland, Trini López Fernández, Living with PCD patient advisory group, Jane S Lucas, Claudia E Kuehni

## Abstract

**Introduction:** Pulmonary exacerbations contribute to disease progression in chronic lung diseases. In a large prospective cohort study, we studied the incidence and predictors of pulmonary exacerbations among persons with primary ciliary dyskinesia (PCD), which can inform follow-up care. We also assessed healthcare use, changes in management, and pathogens during exacerbations.

**Methods:** Participants in the *Living with PCD* study reported increased respiratory symptoms in the past seven days, indicating a pulmonary exacerbation, from June 2020 through May 2022 via online questionnaires. We derived incidence rates and studied predictors of pulmonary exacerbation incidence by fitting multivariable negative binomial regression models.

**Results:** We obtained data from 660 persons (408 adults, 57 adolescents, 195 children) who completed 17,853 follow-up questionnaires (median 17, range 1-84). The 1026 reported exacerbations indicate an incidence rate of 3.1 pulmonary exacerbations per person per year, with minor variation across age groups, but changes over time. Incidence was higher among adult females [incidence rate ratio (IRR) 2.0, 95% confidence interval (CI) 1.4-2.7] and those in whom *Pseudomonas aeruginosa* was isolated (children IRR 1.9, 95% CI 1.1-3.6; adults IRR 1.4, 95% CI 1.0-1.9). Participants saw a health professional during only 185 of 1404 exacerbation weeks (13%). *Pseudomonas aeruginosa* was the pathogen most frequently observed during exacerbations in children (18 of 118 samples, 15%) and adults (132 of 303 samples, 44%).

**Conclusion:** Pulmonary exacerbations are frequent in PCD and heighten the disease burden. Patients for whom targeted management is particularly important include adult females and those who carry *Pseudomonas aeruginosa*.

## Introduction

Pulmonary exacerbations alter disease management and may contribute to disease progression in primary ciliary dyskinesia (PCD) [1]. Impaired mucociliary clearance leads to recurrent infections, driving more frequent exacerbations, which are likely to result in progressive lung damage and faster deterioration of lung function as in other suppurative lung diseases [2–4]. Exacerbations are primarily triggered by bacterial and viral infections, but host and environmental factors such as pollution and chronic inflammation likely increase the risk of exacerbations [4, 5]. Not all patients recover to baseline lung function after an exacerbation [6, 7], which emphasises the importance of effective prevention and timely management.

Knowing which persons with PCD are at higher risk of exacerbation would enable earlier and more intensive treatment and improve clinical follow-up. However, prospective standardised data on pulmonary exacerbations are scarce. While azithromycin reduced respiratory exacerbations by 50% compared to placebo in a randomised controlled trial (RCT) [8]; data on other factors influencing exacerbation incidence are limited [9–11]. Additionally, inconsistent definitions in previous studies have made comparability difficult, with a consensus definition only recently published [1].

Using data from repeated self-reported questionnaires in a prospective cohort study, we estimated the incidence of pulmonary exacerbations in PCD and studied predictors of higher incidence. We also assessed healthcare use, changes in management, and reported pathogens during exacerbations.

## Methods

### Study design and procedure

We studied pulmonary exacerbations using data from the *Living with PCD* study, an international prospective cohort study initiated in May 2020 by people with PCD and the PCD research team at the University of Bern, Switzerland. The study protocol has been published under its previous name COVID-PCD [12]. All people with a confirmed or suspected PCD diagnosis can participate. The initial primary study aim was to monitor health during the coronavirus disease 2019 (COVID-19) pandemic. Participants were invited to take part via patient associations worldwide that advertised the study.

Participants registered online through the study website (pcd.ispm.ch) and participated anonymously. We collected data through online questionnaires available in five languages (English, French, German, Italian, and Spanish) and used the Research Electronic Data Capture (REDCap) database hosted at the Centre for Medical Registries and Data Linkage at the University of Bern, Switzerland [13]. Participants or their caregivers give informed consent when enrolling in the study and can leave the study at any time by informing the study team. The cantonal ethics committee of Bern, Switzerland, has approved the study (study ID: 2020-00830).

### Data collection

We collected data through questionnaires at baseline and through weekly (June 2020 through December 2021) and monthly (January 2022 through May 2022) follow-up questionnaires. The baseline questionnaire covered sociodemographic information, PCD diagnosis, symptoms and clinical manifestations, isolated pathogens, and antibiotic usage based on modules from the FOLLOW-PCD questionnaire [14]. The follow-up questionnaires assessed weekly symptoms, healthcare usage, COVID-19 infections, and isolation of pathogens.

### Inclusion criteria

This analysis included all participants who completed the baseline and at least one follow-up questionnaire between June 2020 and May 2022.

### Pulmonary exacerbations

Weekly questionnaires assessed new symptoms or increase in chronic respiratory symptoms in the previous seven days. We covered all items that constituted an exacerbation according to the BEAT-PCD definition [1]. Supplementary Table S1 lists the elements of the definition and corresponding questions. We labelled any week in which the definition of a pulmonary exacerbation was met as an exacerbation week. Since exacerbations can last longer than one week, we counted consecutive exacerbation weeks as one exacerbation. We only counted a new exacerbation if the participant reported at least one asymptomatic week since the last exacerbation or if no exacerbation week was reported in the last 20 days, assuming that exacerbations usually do not last longer than four weeks. We assessed the duration of an exacerbation as the number of consecutive exacerbation weeks. We checked for outliers and implausible values and excluded participants with an implausible proportion of exacerbation weeks (> 70% of all weeks) reflecting possible misunderstanding of questions.

### Exposures of interest and other variables

We considered variables previously associated with pulmonary exacerbations or severe airway disease in PCD or other suppurative lung diseases as potential predictors of exacerbation incidence: sex, age, age at diagnosis, affected genes, prophylactic antibiotics, regular physiotherapy, physical activity, body mass index (BMI), and environmental tobacco smoke (ETS). As proxies for disease severity we considered bronchiectasis, congenital heart disease, *Pseudomonas aeruginosa* isolation, and forced expiratory volume in the first second (FEV1) [9, 10, 15–17]. All variables were assessed at baseline. Supplementary Table S2 defines the variables.

The follow-up questionnaires assessed social contact behaviour categorised as shielding and reduced contacts, and investigated healthcare use and change of therapies during exacerbations (Supplementary Table S2). We assessed isolated pathogens and positive COVID-19 tests during and outside of exacerbations.

### Statistical analysis

We calculated incidence rates of pulmonary exacerbations per person per year by dividing the number of pulmonary exacerbations by person-years at risk. We defined person-years at risk as the time in years participants were at risk of an exacerbation, assigning 7 days of follow-up to each completed questionnaire and subtracting the time participants already had an exacerbation. We calculated incidence rates stratified by age groups. To assess changes of incidence rate over time, we calculated monthly incidence rates and plotted them together with social contact behaviour (shielding, reduced contacts). We calculated the proportion of exacerbation weeks in all follow-up weeks by dividing the number of exacerbation weeks by the number of completed follow-up questionnaires.

We tested the robustness of the incidence rate calculations in three sensitivity analyses. In the first, we changed the definition of time at risk by not subtracting the time during which participants already had an exacerbation. In the second, we changed the period between two exacerbations to 14 days, and in the third to 30 days, testing the impact of our assumption that exacerbations do not last longer than four weeks.

We studied predictors of higher incidence of pulmonary exacerbations using negative binomial regression analysis with number of exacerbation weeks as outcome and number of completed questionnaires as offset; we chose this model because of the overdispersion in the outcome data. We chose number of exacerbation weeks as outcome to give more weight to longer (i.e. more severe) exacerbations. In this analysis, the response “I don’t know” for congenital heart disease and bronchiectasis was categorised as “no”, while participants with sex “other” and those with missing values for age at diagnosis and physiotherapy were excluded from the analysis. A priori we included sex, age, age at diagnosis, bronchiectasis, congenital heart disease, prophylactic antibiotics, and regular physiotherapy in the model and tested all other predictors in univariable analyses, and subsequently also included *Pseudomonas aeruginosa* isolation since it was strongly associated with incidence. Due to lack of associations in the univariable model, we did not further investigate physical activity, BMI, and ETS. We did not include gene groups and FEV1 in our main model since these variables were only available for some participants. After testing for nonlinear effects, we included age as a quadratic term. We identified an interaction between age and sex in our main model and, to facilitate interpretation, we a posteriori stratified our main model into children (< 14 years) and adults (≥14 years). We chose this cut-off due to the hormonal changes during this period, which influence chronic lung disease outcomes [18], and because parents completed the questionnaires for individuals younger than 14 years. Age-stratified models included the following groups: children: < 7 years, 7-13 years; adolescents/adults: 14-20 years, 21-40 years, 41-60 years, > 60 years.

In subgroups with available data, we analysed effects of FEV1 and affected genes. Two sensitivity analyses tested the robustness of our regression model by using the number of exacerbations instead of weeks of exacerbation as the outcome, and by excluding participants who completed more than ten questionnaires and reported exacerbations in more than 50% of these questionnaires. This reduced the influence of participants with severe disease and many exacerbations. In a post hoc analysis we explored the higher incidence in adult females by testing the hypothesis of disease transmission from child to mother by including “living with a child” as a predictor. We performed all analysis in R statistical software, version 4.4.1 [19].

## Results

We included data from 660 individuals (408 adults, 57 adolescents, 195 children) with at least one follow-up questionnaire completed through May 2022 (Table 1). Most were female (399, 60%) with a median age of 28 years [interquartile range (IQR) 12-45] at registration. Participants came from 49 countries, most from North America (139, 21%), the United Kingdom (136, 21%), and Germany (99, 15%). At registration, 356 of 465 adults and adolescents (77%) and 52 of 195 children (27%) reported bronchiectasis, and 309 (47%) reported prophylactic antibiotics. Participants completed a total of 17,853 follow-up questionnaires between 7 June 2020 and 27 May 2022 (median 17, range 1-84); 40% of questionnaires they received (Table 2, Figure 1A). We excluded 13 follow-up questionnaires completed after May 2022 or with missing information on symptoms.

**Figure 1:**
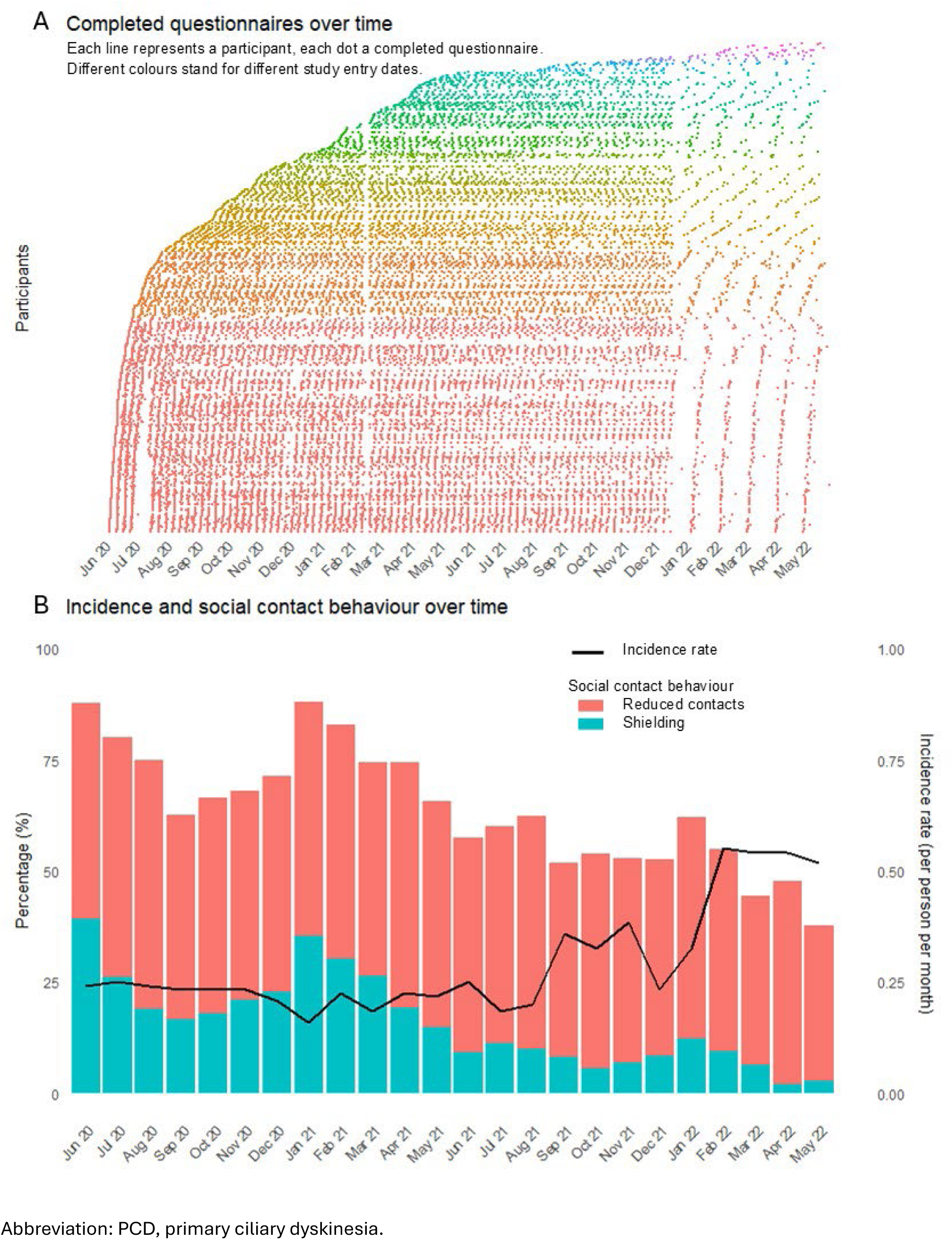
(A) Completed questionnaires and (B) incidence of pulmonary exacerbations and social contact behaviour over time in the *Living with PCD* study (N = 17,853)

**Table 1:** Characteristics of people with primary ciliary dyskinesia participating in the *Living with PCD* study, overall and by age groups (N = 660)

| Characteristics |  | Total<br>N = 660 | Adults/adolescents<br>N = 465 | Children<br>N = 195 |
| --- | --- | --- | --- | --- |
| Sex | Female | 399 (60) | 314 (68) | 85 (44) |
|  | Male | 259 (39) | 149 (32) | 110 (56) |
|  | Other | 2 (0) | 2 (0) | 0 (0) |
| Age | Median, IQR (in y) | 28, 12-45 | 38, 26-50 | 7, 5-11 |
|  | < 7 y | 87 (13) | - | 87 (45) |
|  | 7-13 y | 108 (16) | - | 108 (55) |
|  | 14-20 y | 57 (9) | 57 (12) | - |
|  | 21-40 y | 207 (31) | 207 (45) | - |
|  | 41-60 y | 162 (25) | 162 (35) | - |
|  | > 60 y | 39 (6) | 39 (8) | - |
| Country/region<sup>a</sup> | North America | 139 (21) | 94 (20) | 45 (23) |
|  | United Kingdom | 136 (21) | 97 (21) | 39 (20) |
|  | Germany | 99 (15) | 59 (13) | 40 (21) |
|  | Switzerland | 46 (7) | 36 (8) | 10 (5) |
|  | France | 40 (6) | 27 (6) | 13 (7) |
|  | Italy | 38 (6) | 29 (6) | 9 (5) |
|  | Other European countries | 109 (17) | 86 (18) | 23 (12) |
|  | Other non-European countries | 53 (8) | 37 (8) | 16 (8) |
| Age at diagnosis | Median, IQR | 7, 2-18 | 12, 6-27 | 2, 0-6 |
|  | Missing | 20 (3) | 15 (3) | 5 (3) |
| Congenital heart disease | Yes | 53 (8) | 26 (6) | 27 (14) |
|  | I don't know | 25 (4) | 19 (4) | 6 (3) |
| Bronchiectasis | Yes | 408 (62) | 356 (77) | 52 (27) |
|  | I don't know | 58 (9) | 39 (8) | 19 (10) |
| Prophylactic antibiotics | Yes | 309 (47) | 215 (46) | 94 (48) |
| Regular physiotherapy | Yes | 458 (69) | 296 (64) | 162 (83) |
|  | Missing | 4 (1) | 3 (1) | 1 (1) |
| Gene groups<sup>b</sup> |  | n = 186 | n = 106 | n = 80 |
|  | DS | 117 (63) | 68 (64) | 49 (61) |
|  | DA | 18 (10) | 11 (10) | 7 (9) |
|  | N-DRC | 33 (18) | 17 (16) | 16 (20) |
|  | RS-CC | 16 (9) | 9 (8) | 7 (9) |
|  | other | 2 (1) | 1 (1) | 1 (1) |
| FEV1 |  | n = 497 | n = 371 | n = 126 |
|  | ≥ 60% predicted | 308 (62) | 223 (60) | 85 (67) |
|  | < 60% predicted | 86 (17) | 83 (22) | 3 (2) |
|  | I don't know | 103 (21) | 65 (18) | 38 (30) |
| *Pseudomonas aeruginosa* isolation | Yes | 148 (22) | 131 (28) | 17 (9) |

Abbreviations: DA, dynein assembly. DS, dynein structure. FEV1, Forced expiratory volume in the first second. IQR, interquartile range. N-DRC, microtubular stabilisation/nexin-dynein regulatory complex. PCD, primary ciliary dyskinesia. RS-CC, radial spoke and central complex. y, years. <sup>a</sup> Countries or regions with N≥30 displayed in table, countries with N<30 were categorised into other European or other non-European countries. <sup>b</sup> We grouped affected genes according to their role in cilia structure and function, as done previously (Shoemark et al., 2021). All characteristics are presented as n and column %, unless otherwise stated.

**Table 2:**
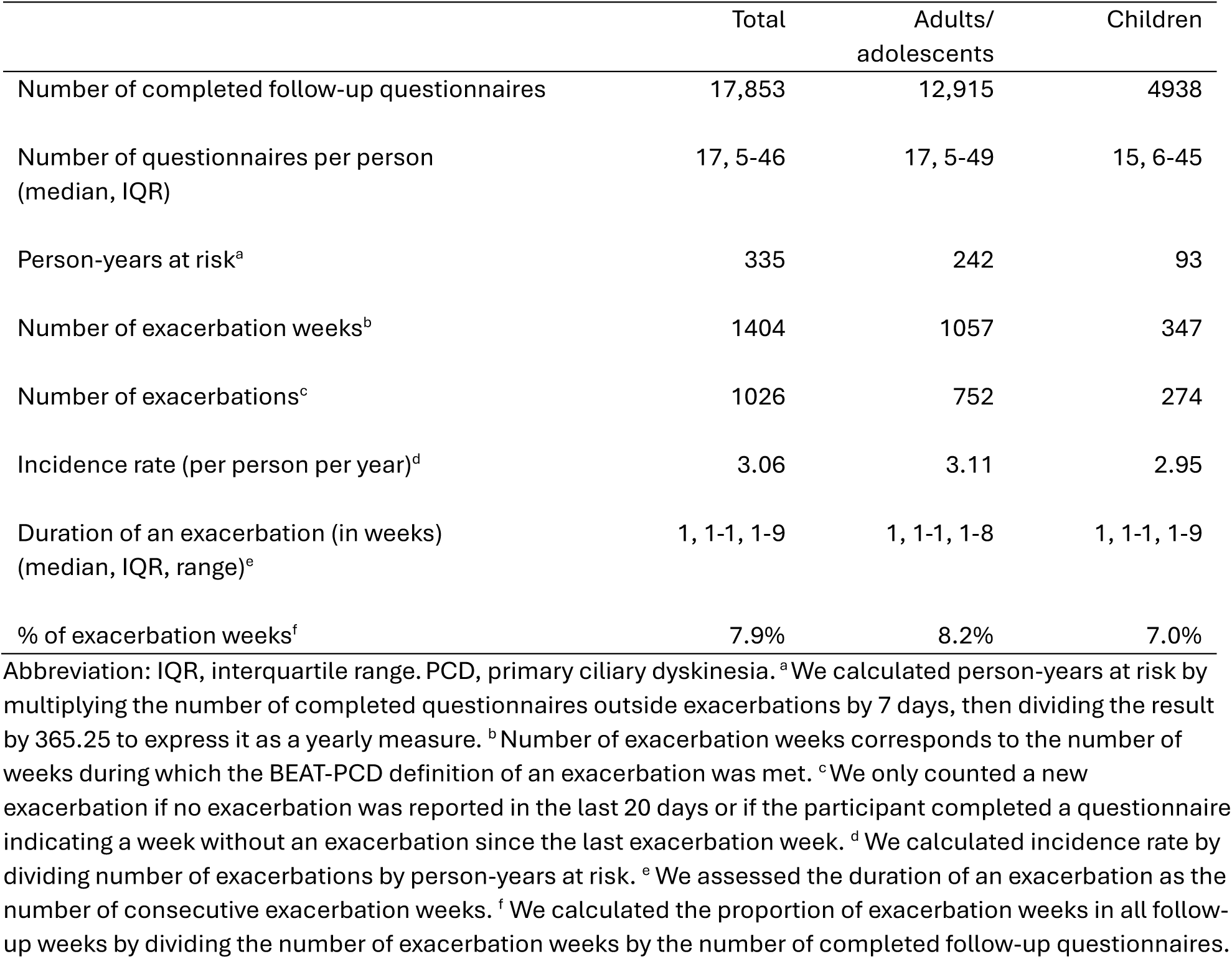
Pulmonary exacerbation incidence and duration in the *Living with PCD* study, by age groups (N = 17,853)

|  | Total | Adults/<br>adolescents | Children |
| --- | --- | --- | --- |
| Number of completed follow-up questionnaires | 17,853 | 12,915 | 4938 |
| Number of questionnaires per person<br>(median, IQR) | 17, 5-46 | 17, 5-49 | 15, 6-45 |
| Person-years at risk<sup>a</sup> | 335 | 242 | 93 |
| Number of exacerbation weeks<sup>b</sup> | 1404 | 1057 | 347 |
| Number of exacerbations<sup>c</sup> | 1026 | 752 | 274 |
| Incidence rate (per person per year)<sup>d</sup> | 3.06 | 3.11 | 2.95 |
| Duration of an exacerbation (in weeks)<br>(median, IQR, range)<sup>e</sup> | 1, 1-1, 1-9 | 1, 1-1, 1-8 | 1, 1-1, 1-9 |
| % of exacerbation weeks<sup>f</sup> | 7.9% | 8.2% | 7.0% |

Abbreviation: IQR, interquartile range. PCD, primary ciliary dyskinesia. <sup>a</sup>We calculated person-years at risk by multiplying the number of completed questionnaires outside exacerbations by 7 days, then dividing the result by 365.25 to express it as a yearly measure. <sup>b</sup>Number of exacerbation weeks corresponds to the number of weeks during which the BEAT-PCD definition of an exacerbation was met. <sup>c</sup>We only counted a new exacerbation if no exacerbation was reported in the last 20 days or if the participant completed a questionnaire indicating a week without an exacerbation since the last exacerbation week. <sup>d</sup>We calculated incidence rate by dividing number of exacerbations by person-years at risk. <sup>e</sup>We assessed the duration of an exacerbation as the number of consecutive exacerbation weeks. <sup>f</sup>We calculated the proportion of exacerbation weeks in all follow-up weeks by dividing the number of exacerbation weeks by the number of completed follow-up questionnaires.

### Incidence of pulmonary exacerbations

Overall incidence was 3.06 pulmonary exacerbations per person per year with only minor variation across age groups (3.11 for adults and adolescents, 2.95 for children). This rate was based on a total of 1026 exacerbations occurring during 335 person-years at risk (Table 2). 235 exacerbations lasted longer than one week, resulting in 1404 of 17,853 follow-up weeks (7.9%) meeting the definition of an exacerbation week.

Exacerbation rates changed over the study period, with reduced social interactions corresponding with lower exacerbation rates independent of season (Figure 1B). Incidence rates ranged between 0.16 and 0.55 per person per month in the two years of follow-up. From spring/summer 2021, we observed an increase in incidence of exacerbations, with an ongoing reduction in shielding behaviour and increased social contacts. Sensitivity analyses with different definitions for exacerbations and time at risk yielded consistent incidence rates (Supplementary Table S3).

### Predictors of higher incidence of pulmonary exacerbations

The fully adjusted multivariable negative binomial regression analysis suggested that people in whom *Pseudomonas aeruginosa* was identified and those taking prophylactic antibiotics had more exacerbations (Table 3). Age, age at diagnosis, and bronchiectasis were not associated with incidence. Since we identified an interaction between sex and age, we stratified the main model by age and created separate models for adults and children. In the adult-only model, the incidence among adult females was twice that of adult males [incidence rate ratio (IRR) 2.0, 95% confidence interval (CI) 1.4-2.7] (Figure 2). Congenital heart disease and *Pseudomonas aeruginosa* isolation were also associated with higher incidence among adults (congenital heart disease: IRR 2.0, 95% CI 1.1-3.7; *Pseudomonas aeruginosa*: IRR 1.4, 95% CI 1.0-1.9). *Pseudomonas aeruginosa* was a predictor for higher incidence also among children (IRR 1.9, 95% CI 1.1-3.6).

**Figure 2:**
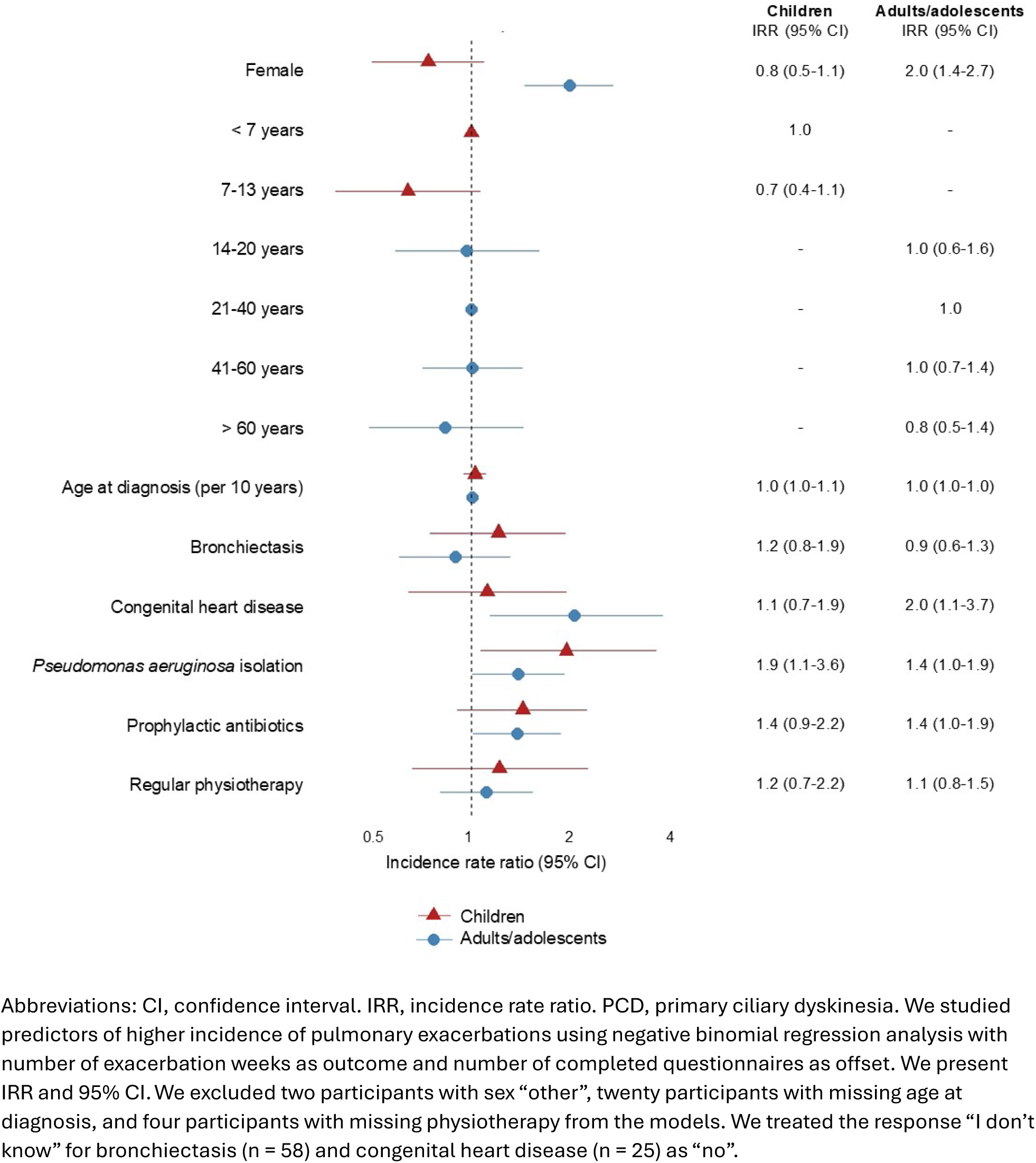
Potential predictors for higher exacerbation incidence in the *Living with PCD* study, by children (n = 189) and adults/adolescents (n = 445)

**Table 3:** Potential predictors for higher exacerbation incidence among people with primary ciliary dyskinesia included in the *Living with PCD* study (N = 660)

|  |  | Unadjusted | p-value | Fully adjusted | p-value |
| --- | --- | --- | --- | --- | --- |
|  |  | n = 660<br>IRR (95% CI) |  | n = 634<br>IRR (95% CI) |  |
| **Sex** (ref.: Male)<sup>a</sup> | Female | 1.4 (1.1-1.8) | 0.003 | 0.9 (0.6-1.3) | 0.478 |
| **Age** | Per year | 1.00 (1.00-1.01) | 0.394 | 1.00 (0.98-1.03) | 0.711 |
| **Age \* age** | Per year<sup>2</sup> | 1.00 (1.00-1.00) | 0.800 | 1.00 (1.00-1.00) | 0.064 |
| **Sex \* age** | Per year | 1.01 (1.00-1.03) | 0.014 | 1.02 (1.00-1.03) | 0.006 |
| **Age at diagnosis**<sup>b</sup> | Per 10 years | 1.04 (0.96-1.1) | 0.272 | 1.1 (0.99-1.2) | 0.062 |
| **Bronchiectasis** (ref.: No)<sup>c</sup> | Yes | 1.3 (1.04-1.7) | 0.019 | 1.01 (0.8-1.4) | 0.953 |
| **Congenital heart disease** (ref.: No)<sup>c</sup> | Yes | 1.4 (0.9-2.0) | 0.144 | 1.6 (1.1-2.4) | 0.026 |
| **P. aeruginosa isolation** (ref.: No) | Yes | 1.7 (1.3-2.1) | <0.001 | 1.5 (1.1-1.9) | 0.004 |
| **Prophylactic antibiotics** (ref.: No) | Yes | 1.6 (1.3-2.0) | <0.001 | 1.4 (1.1-1.7) | 0.011 |
| **Regular physiotherapy** (ref.: No)<sup>b</sup> | Yes | 1.2 (0.95-1.6) | 0.095 | 1.02 (0.8-1.4) | 0.560 |

Abbreviations: CI, confidence interval. IRR, incidence rate ratio. P. aeruginosa, *Pseudomonas aeruginosa*. PCD, primary ciliary dyskinesia. ref, reference. <sup>a</sup>We excluded two participants with sex “other” from the model. <sup>b</sup>We excluded twenty participants with missing age at diagnosis and four participants with missing physiotherapy from the model. <sup>c</sup>We treated the response “I don’t know” for bronchiectasis (n = 58) and congenital heart disease (n = 25) as “no”. We studied predictors of higher incidence of pulmonary exacerbations using negative binomial regression analysis with number of exacerbation weeks as outcome and number of completed questionnaires as offset. The fully adjusted model included all listed predictors. We present IRR and 95% CI.

In subgroups, we also analysed effects of FEV1 and affected genes on pulmonary exacerbation incidence but found no associations (Supplementary Table S4). To explore the higher incidence in adult females, we conducted a post hoc analysis that included “living with a child” as a predictor, but found no association. In sensitivity analyses, the findings from our main model stayed consistent if we used number of exacerbations as outcome instead of exacerbation weeks or if we excluded participants with very frequent exacerbations (Supplementary Table S4).

### Healthcare use, treatment changes, and pathogens

Participants contacted a healthcare professional by phone during 758 of 1404 exacerbation weeks (54%) and had an appointment with a healthcare professional during 185 (13%) of exacerbation weeks. Participants often managed the exacerbation themselves by changing therapies (1166 exacerbation weeks, 83%), such as starting antibiotic treatment (829 weeks, 59%), increasing physiotherapy (630 weeks, 45%), or increasing inhaler use (455 weeks, 32%) (Table 4A).

**Table 4:** (A) Healthcare use and change of therapies during exacerbations, and (B) reported pathogens and COVID-19 samples during and outside exacerbations in the *Living with PCD* study, by adults/adolescents and children.

| During exacerbations | Overall<br>N = 1404 | Adults/<br>adolescents<br>N = 1057 | Children<br>N = 347 |
| --- | --- | --- | --- |
| **Healthcare use** | **789 (56)** | **560 (53)** | **229 (66)** |
| Contact with health professional by phone due to symptoms | 758 (54) | 545 (52) | 213 (61) |
| Appointment with health professional due to symptoms | 185 (13) | 124 (12) | 61 (18) |
| **Change of therapies** | **1166 (83)** | **861 (81)** | **305 (89)** |
| - Start of antibiotic treatment | 829 (59) | 609 (58) | 220 (63) |
| - More physiotherapy | 630 (45) | 470 (44) | 160 (46) |
| - More frequent use of inhaler | 455 (32) | 382 (36) | 73 (21) |
| - More nasal rinsing | 368 (26) | 302 (29) | 66 (19) |
| - More physical activity | 109 (8) | 83 (8) | 26 (7) |

(B) Pathogens and COVID-19
|  | During exacerbations |  | Outside exacerbations |  |
| --- | --- | --- | --- | --- |
|  | Adults/<br>adolescents | Children | Adults/<br>adolescents | Children |
| **Pathogens** |  |  |  |  |
| Number of samples | N = 303 | N = 118 | N = 517 | N = 401 |
| Pathogens detected: |  |  |  |  |
| *Pseudomonas aeruginosa* | 132 (44) | 18 (15) | 153 (30) | 28 (7) |
| *Haemophilus influenzae* | 14 (5) | 12 (10) | 30 (6) | 36 (9) |
| MSSA | 9 (3) | 10 (8) | 33 (6) | 33 (8) |
| MRSA | 12 (4) | 3 (3) | 8 (2) | 4 (1) |
| *Streptococcus pneumoniae* | 8 (3) | 8 (7) | 14 (3) | 14 (3) |
| *Candida species* | 17 (6) | 10 (8) | 32 (6) | 15 (4) |
| *Aspergillus fumigatus* | 11 (4) | 13 (11) | 28 (5) | 11 (3) |
| **COVID-19** |  |  |  |  |
| Number of COVID-19 samples | N = 290 | N = 116 | N = 1231 | N = 524 |
| COVID-19 positive | 24 (8) | 21 (18) | 26 (2) | 22 (4) |

Abbreviations: COVID-19, coronavirus disease 2019. MSSA, methicillin-susceptible staphylococcus aureus. MRSA, methicillin-resistant staphylococcus aureus. PCD, primary ciliary dyskinesia. All characteristics are presented as n and column %.

During exacerbations, *Pseudomonas aeruginosa* was the pathogen most frequently identified in both adults (132 of 303 samples, 44%) and children (18 of 118 samples, 15%). Other pathogens were rare in adults, while in children *Aspergillus fumigatus* (13 of 118 samples, 11%) and *Haemophilus influenzae* (12 of 118 samples, 10%) were found with frequency similar to *Pseudomonas aeruginosa* (Table 4B). Outside of exacerbations, *Pseudomonas aeruginosa* remained the most common pathogen in adults, while in children, *Haemophilus influenzae* was slightly more prevalent. During the study period, participants reported 2161 COVID-19 tests of which 93 (4%) were positive; half of the positive samples (45, 48%) were reported during an exacerbation.

## Discussion

Using prospective patient-reported data collected regularly over a two-year period, we estimated the incidence rate of pulmonary exacerbations among people with PCD to be 3.1 per person per year with minimal variation between age groups but with changes over time. *Pseudomonas aeruginosa* was frequently detected during exacerbations in children and adults and incidence of exacerbations was higher among adult females and participants from whom *Pseudomonas aeruginosa* was isolated. Participants often managed exacerbations themselves without visiting healthcare professionals.

Our study is one of few to use prospective data on the incidence of pulmonary exacerbations. We collected weekly information on symptoms and healthcare use, and our comprehensive baseline questionnaire allowed investigating several predictors. Our study preserved participant anonymity, thus, self-reported data could not be verified with clinical records. However, the standard definition of pulmonary exacerbation in PCD used in research is based on patient-reported measures anyway [1]. The completion rate of weekly questionnaires (40%) may be a limitation in estimating the incidence rate since it may have resulted in under-reporting (if not all pulmonary exacerbations were recorded) and over-reporting (if participants were more likely to complete questionnaires when symptomatic). We found that participants who completed more questionnaires had lower incidence rates, which favours an overestimation of the true incidence rate in our study (data not shown). In addition, the changes in pathogen incidence during the COVID-19 pandemic due to reduced social contacts and mask-wearing might affect the generalisability of our results to periods outside the pandemic. The frequency of pathogens transmitted via droplet infection, such as *Haemophilus influenzae*, was reduced during the pandemic [20], which we also observed in our study (data not shown).

We observed similar incidence rates in our study to those reported in the placebo group of the RCT comparing azithromycin to placebo, where all respiratory exacerbations were recorded [8]. However, as per inclusion criteria of the RCT, none of the patients were colonised with *Pseudomonas aeruginosa* and none received prophylactic antibiotics [8] so they had a lower disease severity than our participants.

Adult females in our study experienced more exacerbations than males. This is consistent with prior findings in PCD showing higher respiratory morbidity in females [15] and it mirrors other respiratory diseases, such as cystic fibrosis, in which females experience more exacerbations after puberty than males, while the reverse trend is observed in children [21]. In these diseases, hormonal influences on the airway surface liquid have been proposed as a potential mechanism. Higher oestrogen levels after puberty may compromise airway surface liquid dynamics needed for effective mucociliary clearance [22]; however, mucociliary clearance is already compromised in females with PCD. Another consideration is that females may be more self-aware of their symptoms, which could lead to increased reporting of symptoms and more proactive health-seeking behaviour. We also explored whether caring for a child, to which mothers usually have closer contact than fathers, contributes to the explanation. We were not able to confirm this hypothesis; however, our data set for analysis was rather small and lacked power. Genetic, morphological, immunological, environmental and lifestyle-related factors can also contribute to sex differences.

*Pseudomonas aeruginosa* isolation was strongly associated in our analysis with pulmonary exacerbation incidence, which is consistent with previous studies linking *Pseudomonas aeruginosa* colonisation with poor lung outcomes [9, 15, 23]. However, the presence of *Pseudomonas aeruginosa* may be less a predictor of exacerbations than it is a marker of advanced lung disease.

Contrary to intuition, our main model found no significant association between bronchiectasis and pulmonary exacerbations. We cannot know if the effect of bronchiectasis may have been obscured by unmeasured confounders. Finally, we found an association of prophylactic antibiotics with more exacerbations, which likely reflects confounding by indication. Patients receiving prophylactic antibiotics have more severe disease, which may explain why their exacerbation rates remain high despite treatment. The lack of seasonality in the exacerbation incidence combined with the dependence on social contact behaviour suggests that the anecdotally reported higher incidence in the winter months outside the pandemic may be due to changed social contact behaviour rather than environmental factors such as temperature, humidity, or heating use. This complexity of causal relationships between infections, lung damage, therapeutic interventions, and environmental factors highlights the need for further prospective studies to better understand the differentiating factors leading to exacerbations.

The consensus definition for pulmonary exacerbations [1] has not yet been validated, and it remains uncertain whether it fully captures true exacerbations. It does not distinguish between more and less severe exacerbations and does not consider the duration of symptoms. In young children, exacerbations are often due to colds—i.e. viral infections that are also common in this age group in the general population. In addition, it may be difficult for people with severe illness who do not recover to baseline to accurately report increased symptoms because they cannot clearly distinguish between periods of exacerbation and stability. The definition needs to be validated to ensure the significance of future research on pulmonary exacerbations.

In conclusion, this study confirms that pulmonary exacerbations are frequent in PCD and contribute to high disease morbidity. Adult females and those in whom *Pseudomonas aeruginosa* can be identified are at particular risk, highlighting the importance of targeted management in these groups.

## Data availability

*Living with PCD* data are available upon reasonable request by contacting Claudia Kuehni.

## Data Availability

*Living with PCD* data are available upon reasonable request by contacting Claudia Kuehni.

## Acknowledgements

We thank all participants and their families, and we thank the PCD support groups and physicians who advertised the study. We thank our collaborators who helped set up the *Living with PCD* study: Cristina Ardura, Yin Ting Lam, Christina Mallet, Helena Koppe, Dominique Rubi from the University of Bern, and Amanda Harris from the University Hospital Southampton. We thank Ben Spycher for his statistical advice and Christopher Ritter for his editorial work and guidance on our manuscript. To improve the readability and clarity of this manuscript, the authors also incorporated suggestions from ChatGPT 4o, DeepL, and Grammarly. After using these tools, the authors reviewed and edited the content as needed and take full responsibility for the content of this work.

## Authors’ roles

LD Schreck, M Goutaki, ESL Pedersen, and CE Kuehni made substantial contributions to the study design. LD Schreck, M Goutaki, ESL Pedersen, F Copeland, T López Fernández, JS Lucas, and CE Kuehni made substantial contributions to the study concept. LD Schreck, M Goutaki, ESL Pedersen, F Copeland, T López Fernández, JS Lucas, and CE Kuehni made substantial contribution to the interpretation of the data. LD Schreck analysed data and drafted the manuscript. LD Schreck, M Goutaki, ESL Pedersen, F Copeland, T López Fernández, JS Lucas, and CE Kuehni critically revised and approved the manuscript.

## Online display item

**Supplementary Table S1:**
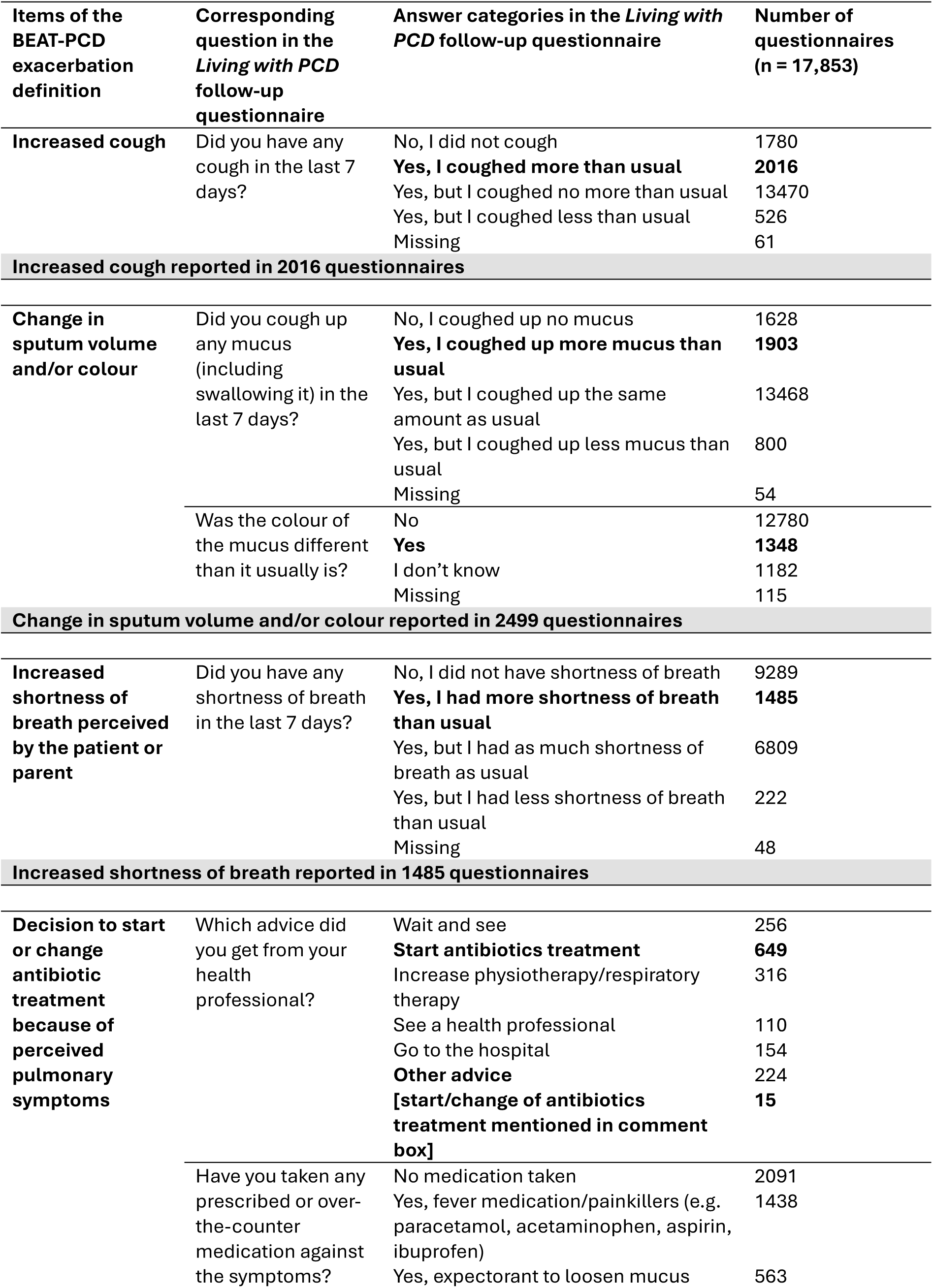
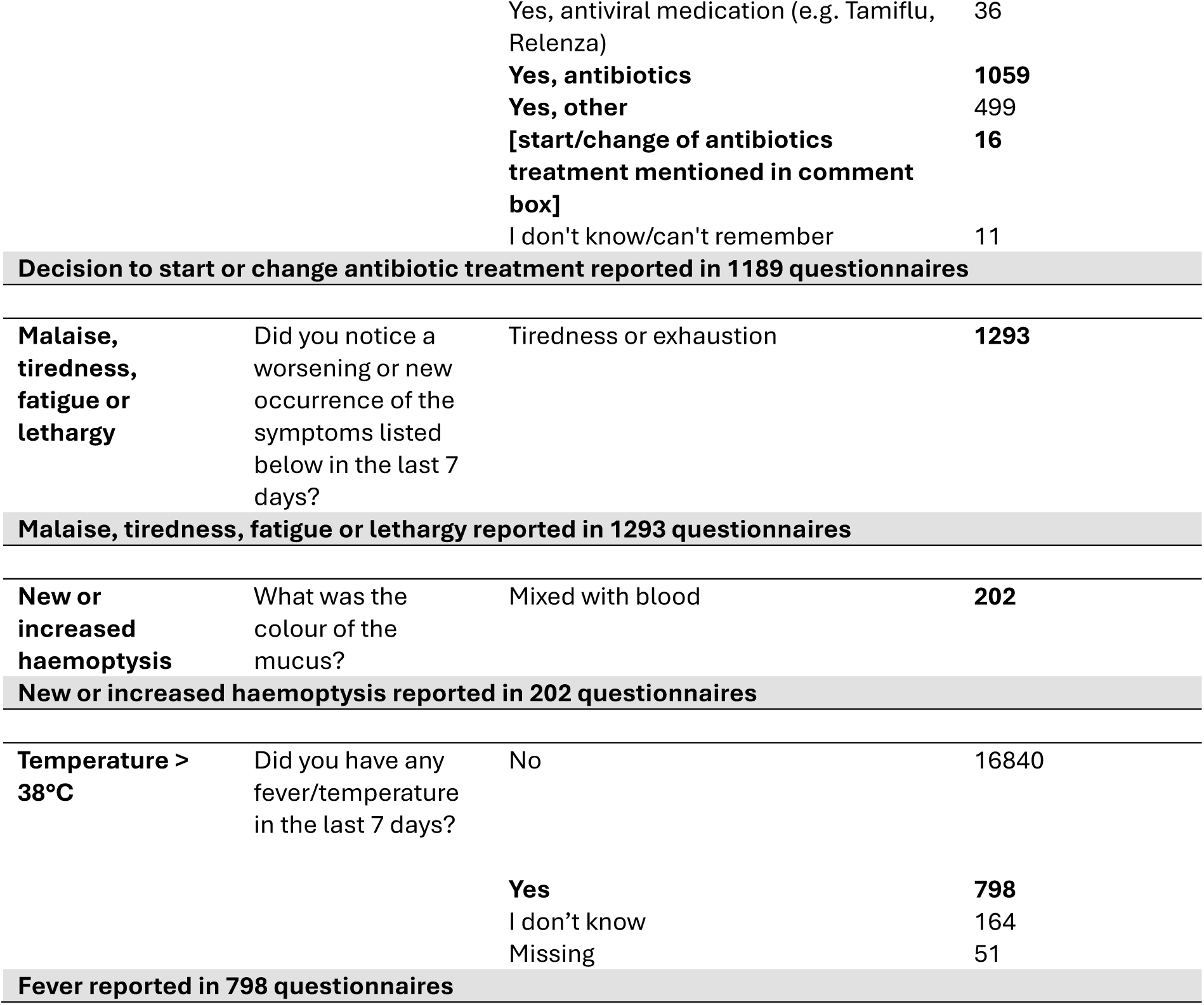
Overview of the items of the BEAT-PCD exacerbation definition with corresponding questions and answer categories in the *Living with PCD* follow-up questionnaires, including the number of questionnaires in which the answer category was ticked by the participants (n = 17,853)

**Supplementary Table S2:** Potential predictors for higher exacerbation incidence and other variables of interest in the *Living with PCD* study.

| Variable | Definition | Questionnaire |
| --- | --- | --- |
| **Predictors** |  |  |
| **Sex** | We categorised sex as “male”, “female”, or “other”. For the regression analysis, we excluded participants with sex “other”. | Baseline |
| **Age** | We used age at registration and included it either as a continuous variable or categorised into age groups; < 7 years, 7-13 years, 14-20 years, 21-40 years, 41-60 years, > 60 years. | Baseline |
| **Country/region** | We categorised countries or regions with N<30 into other European or other non-European countries. | Baseline |
| **Age at diagnosis** | We assessed age at PCD diagnosis, included as continuous variable. We excluded participants with missing age at diagnosis from the regression models. | Baseline |
| **Congenital heart disease** | We categorised the presence of congenital heart disease as “yes”, “no”, or “I don’t know”, with the latter grouped with “no” for regression analysis. | Baseline |
| **Bronchiectasis** | We categorised the presence of bronchiectasis as “yes”, “no”, or “I don’t know”, with the latter grouped with “no” for regression analysis. | Baseline |
| **Prophylactic antibiotics** | We assessed the regular use of antibiotics to prevent bacterial infections and categorised it into “yes” and “no”. | Baseline |
| **Regular physiotherapy** | We assessed the frequency of respiratory physiotherapy/airway clearance in the last month and categorised it into *regular* (several times a week, daily, twice daily or more) and *irregular* (less than once a week/only when I was not well, never). We excluded participants with missing physiotherapy from the regression analysis. | Baseline |
| **Gene groups** | We grouped affected genes according to their role in cilia structure and function (dynein assembly, dynein structure, microtubular stabilisation/nexin-dynein regulatory complex, radial spoke and central complex, other), as done previously (Shoemark et al., 2021). We excluded the group “other” from the regression analysis. | Baseline |
| **FEV1** | We categorised FEV1 as $\geq 60\%$ predicted, $< 60\%$ predicted, and “I don’t know”, with the latter and missing values excluded from the regression analysis. | Baseline |
| ***Pseudomonas aeruginosa* isolation** | We classified participants as “*Pseudomonas aeruginosa* isolated” if they tested positive for *Pseudomonas aeruginosa* in the 12 months before study enrolment. | Baseline |
| **Social contact behaviour** |  |  |
| **Reduced contacts** | We defined reduced contacts if they reported having contact with less than 10 people in the past week but were not shielding. | Follow-up |
| **Shielding** | We defined shielding if participants reported no contact with anyone in the last week or only with those they live with. | Follow-up |
| **Healthcare use** |  |  |
| **Contact with health professional by phone** | We defined contact with a health professional by phone if participants contacted any health professional due to increased symptoms in the last 7 days. | Follow-up |
| **Appointment with health professional** | We defined an appointment with a health professional if they saw any health professional due to increased symptoms in the last 7 days. | Follow-up |
| **Change of therapies** | We defined change of therapies if participants increased or changed any of their usual therapies in the last 7 days. | Follow-up |

**Supplementary Table S3:** Incidence rate estimated by the main analysis, compared to results obtained by alternative approaches to modelling (sensitivity analyses) with a) an alternative calculation of time at risk (including all completed questionnaires) and with a different time span of b) 30 days and c) 14 days between exacerbations, in the *Living with PCD* study.

|  | Main model | Sensitivity analyses |  |  |
| --- | --- | --- | --- | --- |
|  | Total | a<br>Total | b<br>Total | c<br>Total |
| Number of completed follow-up questionnaires | **17,853** | 17,853 | 17,853 | 17,853 |
| Person-years (at risk) | **335** | **342** | 335 | 335 |
| Number of exacerbations | **1026** | 1026 | **990** | **1066** |
| Incidence rate (per person per year) | **3.06** | 3.00 | 2.96 | 3.18 |

Abbreviation: PCD, primary ciliary dyskinesia.

**Supplementary Table S4:** Subgroup, post hoc and sensitivity analyses: Incidence rate ratios of pulmonary exacerbations from a negative binomial regression analysis among people with primary ciliary dyskinesia included in the *Living with PCD* study with a) FEV1 included as a predictor (N = 379), b) gene groups included as a predictor (N = 182), c) “living with a child” included as a predictor in a model which only included adult females (N = 300), d) number of exacerbations as outcome (N = 638), and e) people who reported exacerbations in more than 50% of questionnaires excluded (N = 633).

|  |  | **a**<br>FEV1<br>included as<br>a predictor<br>N = 375<br>IRR (95% CI) | **b**<br>Gene<br>groups<br>included<br>N = 182<br>IRR (95%<br>CI) | **c**<br>“Living with a<br>child”<br>included,<br>only adult<br>females<br>N = 298<br>IRR (95% CI) | **d**<br>Exacerbation<br>as outcome<br>N = 634<br>IRR (95% CI) | **e**<br>People with<br>exacerbation<br>in > 50% of<br>questionnaires<br>excluded<br>N = 629<br>IRR (95% CI) |
| --- | --- | --- | --- | --- | --- | --- |
| **Sex** (ref.: Male)<sup>a</sup> | Female | 0.7 (0.4-1.3) | 0.8 (0.5-1.5) |  | 0.9 (0.6-1.4) | 0.9 (0.6-1.4) |
| **Age** | Per y | 1.0 (1.0-1.1) | 1.0 (1.0-1.0) | 1.0 (1.0-1.1) | 1.0 (1.0-1.0) | 1.0 (1.0-1.0) |
| **Age \* age** | Per y<sup>2</sup> | 1.0 (1.0-1.0) | 1.0 (1.0-1.0) | 1.0 (1.0-1.0) | 1.0 (1.0-1.0) | 1.0 (1.0-1.0) |
| **Sex \* age** (ref.: Male) | Per y | 1.0 (1.0-1.0) | 1.0 (1.0-1.0) |  | 1.0 (1.0-1.0) | 1.0 (1.0-1.0) |
| **Age at diagnosis**<sup>a</sup> | Per 10 y | 1.2 (1.1-1.4) | 1.0 (0.8-1.2) | 1.2 (1.0-1.3) | 1.1 (1.0-1.2) | 1.1 (1.0-1.2) |
| **Bronchiectasis** (ref.: No)<sup>b</sup> | Yes | 0.9 (0.6-1.2) | 1.0 (0.6-1.7) | 1.2 (0.7-1.8) | 1.0 (0.8-1.3) | 1.1 (0.8-1.4) |
| **Congenital heart disease** (ref.: No)<sup>b</sup> | Yes | 1.6 (1.0-2.8) | 1.8 (1.0-3.1) | 2.2 (1.1-4.8) | 1.4 (1.0-2.0) | 1.5 (1.0-2.2) |
| **P. aeruginosa isolation** (ref.: No) | Yes | 1.3 (1.0-1.8) | 2.0 (1.3-3.2) | 1.5 (1.1-2.2) | 1.4 (1.1-1.7) | 1.4 (1.1-1.9) |
| **Prophylactic antibiotics** (ref.: No) | Yes | 1.3 (1.0-1.8) | 1.1 (0.7-1.6) | 1.3 (0.9-1.8) | 1.2 (0.9-1.5) | 1.2 (1.0-1.6) |
| **Regular physiotherapy** (ref.: No)<sup>a</sup> | Yes | 1.1 (0.8-1.6) | 1.4 (0.8-2.4) | 0.8 (0.6-1.2) | 1.1 (0.9-1.4) | 1.3 (1.0-1.6) |
| **Gene groups** (ref.: DS)<sup>a</sup> | DA |  | 1.0 (0.5-1.9) |  |  |  |
|  | N-DRC |  | 0.7 (0.4-1.1) |  |  |  |
|  | RS-CC |  | 1.3 (0.7-2.5) |  |  |  |
| **FEV1** (ref.: ≥ 60%)<sup>a</sup> | < 60% | 1.3 (0.9-1.9) |  |  |  |  |
| **Living with a child** (ref.: No) | Yes |  |  | 1.1 (0.7-1.8) |  |  |

Abbreviations: DA, dynein assembly. DS, dynein structure. CI, confidence interval. FEV1, Forced expiratory volume in the first second. IRR, incidence rate ratio. N-DRC, microtubular stabilisation/nexin-dynein regulatory complex. P. aeruginosa, *Pseudomonas aeruginosa*. PCD, primary ciliary dyskinesia. ref, reference. RS-CC, radial spoke and central complex. y, year. <sup>a</sup> We excluded participants with sex “other”, missing age at diagnosis, missing physiotherapy, gene group “other”, and unknown FEV1 from the respective models. <sup>b</sup> We treated the response “I don’t know” for bronchiectasis (n = 58) and congenital heart disease (n = 25) as “no”. We present IRR and 95% CI.

